# Prediction of Coronary Artery Disease Using Urinary Proteomics

**DOI:** 10.1101/2023.02.09.23285646

**Authors:** Dongmei Wei, Jesus D. Melgarejo, Lucas Van Aelst, Thomas Vanassche, Peter Verhamme, Stefan Janssens, Karlheinz Peter, Zhen-Yu Zhang

## Abstract

**Aims:** Coronary artery disease (CAD) is multifactorial, caused by complex pathophysiology, and contributes to a high burden of mortality worldwide. Urinary proteomic analyses may help to identify predictive biomarkers and provide insights into the pathogenesis of CAD.

**Methods:** Urinary proteome was analyzed in 965 participants using capillary electrophoresis coupled with mass spectrometry. A proteomic classifier was developed in a discovery cohort with 36 individuals with CAD and 36 matched controls using the support vector machine. The classifier was tested in a validation cohort with 115 individuals who progressed to CAD and 778 controls and compared with two previously developed CAD-associated classifiers, CAD238 and ACSP75. The Framingham risk score was available in 737 participants. Bioinformatics analysis was performed based on the CAD-associated peptides.

**Results:** The novel proteomic classifier was comprised of 160 urinary peptides, mainly related to collagen turnover, lipid metabolism, and inflammation. In the validation cohort, the classifier provided an AUC of 0.82 (95% CI: 0.78-0.87) for the CAD prediction in 8 years, superior to CAD238 (AUC: 0.71, 95% CI: 0.66-0.77) and ACSP75 (AUC: 0.53, 95% CI: 0.47-0.60). On top of CAD238 and ACSP75, the addition of the novel classifier improved the AUC to 0.84 (95% CI: 0.80-0.89). In a multivariable Cox model, a 1-SD increment in the novel classifier was associated with CAD (HR: 1.54, 95% CI: 1.26-1.89, P<0.0001) higher risk of CAD. The new classifier further improved the risk reclassification of CAD on top of the Framingham risk score (net reclassification index: 0.61, 95% CI: 0.25-0.95, P=0.001).

**Conclusions:** A novel urinary proteomic classifier related to collagen metabolism, lipids, and inflammation showed potential for the risk prediction of CAD. Urinary proteome provides an alternative approach to personalized prevention.

**Lay summary:**

- A biomarker that can predict coronary artery disease (CAD) is in urgently need.
- We developed and validated a urinary proteomic classifier for the prediction of CAD.
- The proteomic classifier involved in atherosclerosis improved the risk reclassification on top of the clinical risk score.

## Introduction

Coronary artery disease (CAD) is the most common heart disease affecting 126.5 million people and a leading cause of mortality, responsible for an estimated 8.9 million deaths worldwide in 2017.^1^ Despite the advances in the management of modifiable risk factors, residual risk remains.^2^ Given the considerable number of patients with CAD and the growing economic burden it causes,^1^ the need for targeted intervention strategies is urgent. The development of targeted treatments requires insightful inputs into the mechanisms and biomarkers for CAD. CAD can progress asymptomatically, thus biomarkers that can detect the insidious ongoing pathophysiological process prior to clinical events and provide prognostic value are particularly in great demand.

Although blood is a common source of biomarker discovery for cardiovascular diseases,^3^ urine, which can be collected noninvasively and easily, is another reservoir of biomarkers.^4^ Proteins leaking or being secreted from multiple organs, including the cardiovascular system, can enter into the circulation and filter through the kidneys into the urine.^4, 5^ Therefore, a comprehensive urinary proteomics analysis can reflect systemic disease-associated changes, holds the promise to identify predictive biomarkers and uncover the mechanisms for cardiovascular diseases. We have previously reported urinary proteomics biomarkers for arterial stiffness,^6^ vascular calcification,^7^ and heart failure.^8, 9^ Previous multidimensional urinary proteomic biomarkers have also shown the potential for the detection and prediction of CAD.^10-13^ For instance, a panel of 238 urinary peptides, CAD238, provided an AUC of 0.87 for the detection of CAD in 138 samples from 71 CAD patients and 67 healthy controls.^10, 11^ ACSP75 was developed for the prediction of acute coronary syndrome with an AUC of 0.66 in 42 cases and 42 controls.^13^ Given the potential of urinary proteome analysis, there might be room for improvement of the classifier because the performance of a classifier may not be maintained when generalizing to a large cohort. Thus, this study aimed to develop a new urinary proteomic classifier for the prediction of CAD endpoints and validate its prognostic value in 893 individuals. Moreover, we hypothesized that urinary peptides discriminating CAD could provide insights into the pathological processes of CAD at an early stage, thus we comprehensively described the pathways reflected by the identified urinary proteomic biomarkers with bioinformatics approaches.

## Methods

### Study design and participants

The study was a retrospective longitudinal study, comprised of a discovery cohort and a validation cohort (Figure 1). The discovery cohort consisted of 36 cases and 36 matched controls from a population-based study, the Flemish Study on Environment, Genes, and Health Outcomes (FLEMENGHO).^7^ Cases were individuals asymptomatic for CAD at baseline but progressed to CAD during a median of 8.3-year (interquartile [IQR]: 5.5-9.8) follow-up. CAD was defined as myocardial infarction, acute coronary syndrome, coronary artery bypass graft, percutaneous transluminal coronary angioplasty, any fatal ischemic heart disease. Cases and controls with an averaged estimated glomerular filtration rate of 80.5 (IQR: 71.0-89.3) ml/min/1.73m^2^ were matched for sex, age, history of hypertension, antihypertensive treatment, and total cholesterol. The validation cohort was comprised of 893 individuals (115 with and 778 without CAD) from the FLEMENGHO study (35 with and 702 without CAD) or extracted from the Human Urinary Proteome Database (80 with and 76 without CAD), consisting of more than 85000 samples from various clinical and research centers.^14^ All the latter datasets were fully anonymized and previously published, with respective references provided below. Demographic and clinical characteristics from extracted datasets included sex, age, hypertension, diabetes, office blood pressure, and estimated glomerular filtration rate (eGFR). The second use of FLEMENGHO study data (B32220083510) was approved by the University of Leuven Ethics Committee and participants provided written informed consent. Datasets extracted from the Human Urinary Proteome Database were previously published and relevant studies were conducted in compliance with the Helsinki declaration for research in humans and received an approval from the responsible review boards.^13, 15-17^ CAD endpoint was defined as myocardial infarction, acute coronary syndrome, new-onset angina pectoris, ischemic cardiomyopathy, and coronary revascularization.

**Figure 1.**
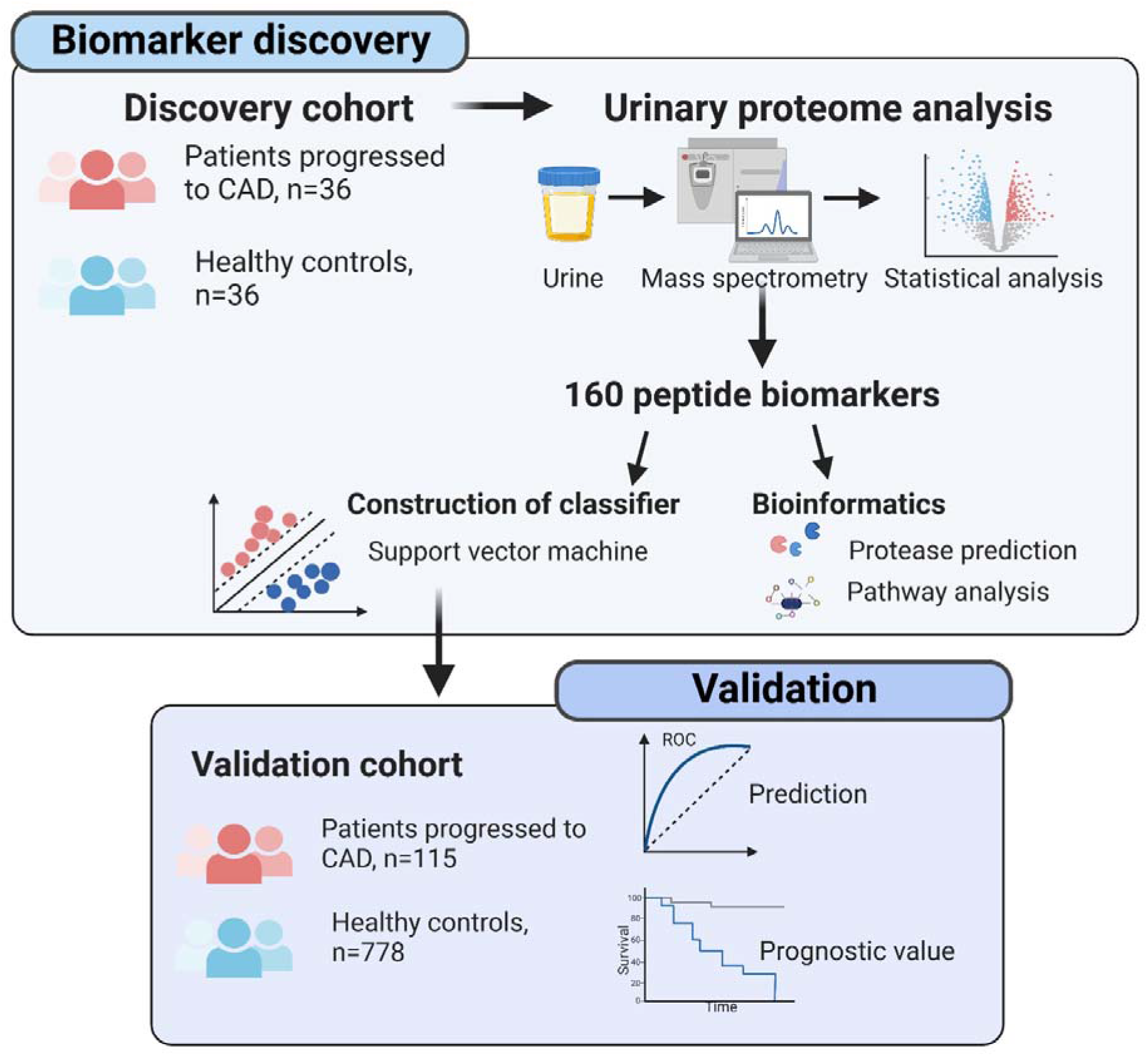
The schematic diagram of study design. In the biomarker discovery phase, urinary proteome analysis was performed on 36 patients who progressed to CAD and 36 matched controls. A total of 160 urinary peptides were identified to be significantly different between cases and controls and their biological function was elucidated by bioinformatic analysis. Simultaneously, 160 peptides were used to construct a classifier for the discrimination of CAD by the supervised machine learning method, and the predictive performance and prognostic value were evaluated by an independent validation cohort. CAD, coronary artery disease.

### Urinary proteome analysis

Urinary proteome analysis was performed with a P/ACE MDQ capillary electrophoresis system (Beckman Coulter, Fullerton, CA) coupled to a micrOTOF MS (Bruker Daltonics, Bremen, Germany). MosaFinder software was used to process mass spectral data and to generate a raw list of peptides or small proteins before being annotated according to prior sequenced peptides from the Human Urinary Proteome Database.^14^ Peptide intensities were normalized using 29 collagen peptides, serving as an internal standards, being in general not affected by the disease, to assure comparability between different datasets.^18^ Further information on sample preparation, data processing, and sequencing of the peptides was described elsewhere.^9, 19, 20^

### Classifier construction

Distinct peptides between individuals who progressed to reach an CAD endpoint and controls were identified in the discovery cohort. Urinary sequenced peptides identified in ≥ 30% of either cases or controls were analyzed. Peptide abundances was compared using the non-parametric Wilcoxon test, followed by adjustment for multiple testing. Peptides were considered statistically significant when the P-value was less than 0.05 after adjustment for multiple testing by Benjamini-Hochberg correction. The statistical validity of significant peptides was further confirmed by permutation analysis that randomly excluded 30% of the samples with 10 times repetition. Peptides with a nominal P-value of <0.05 in more than 50% of the permutation analyses were considered for further analysis. The support vector machine (SVM), a supervised machine learning algorithm, was applied to construct a classifier for the discrimination of CAD using the significant peptides. Model construction and parameters tuning were performed with the MosaCluster software.^21^ The model ‘s generalization was examined by take-one-out cross-validation.

### Statistical analysis

Statistical analysis was conducted with SAS software, version 9.4 (SAS Institute, Cary, NC, USA). Means and percentages were compared using a t-test or analysis of variance (ANOVA) test, or Fisher ‘s test as appropriate. Statistical significance was considered as a two-sided P value of 0.05. Time-dependent receiver operating characteristic (ROC) curves and the areas under the ROC curves (AUC) were used to estimate the predictive capacity of the new classifier for incident CAD at 3, 5, and 8 years. The predictive capacity of previously developed classifiers, CAD238 and ACSP75 were assessed as comparisons.^10-13^ The AUC estimate and 95% confidence interval (CI) for each classifier were calculated using the PHREG procedure to fit the Cox regression model in SAS. Moreover, whether the addition of the urinary proteomic classifier could improve risk reclassification for CAD on top of the Framingham risk score was further evaluated. The Framingham risk score was calculated based on common clinical risk factors, including sex, age, systolic blood pressure, smoking, diabetes, treatment of hypertension, total cholesterol, and high-density lipoprotein cholesterol.^22^ The improvement in risk reclassification was evaluated by net reclassification index (NRI) and integrated discrimination index (IDI). P value and 95% confidence interval (CI) of NRI and IDI were estimated by 500 times bootstrap. The prognostic value of the new classifier was assessed by multivariable Cox proportional hazard models. Model 1 was adjusted for covariates, including sex, age, mean arterial blood pressure, and diabetes. Model 2 was additionally adjusted for eGFR. Hazard ratio (HR) and 95% CI was estimated for per-SD increment in a classifier score or using the bottom quartile of a classifier score as a reference.

### Bioinformatics analysis

To better interpret the underlying mechanisms between identified urinary peptides and CAD, the proteolysis processes that produced the polypeptides were considered. The potential proteases were predicted based on the N- and C-terminal cleavage sites of peptides by the Proteasix Knowledge Base (http://www.proteasix.org).^23^ With the observed prediction mode, the cleavage sites were matched based on the literature. Subsequently, the parental proteins of the peptides, together with the predicted proteases were submitted for pathway enrichment analysis to elucidate their biological functions. The pathway analysis was performed via the ClueGO plug-in (v 2.5.7) of Cytoscape v 3.7.2 using the Reactome pathway database (updated on 8, May 2020).^24, 25^ The minimum number of proteins to enrich a pathway was 3 and the minimum enrichment ratio was 4%. The significant threshold of P-values corrected by the Bonferroni step-down was 0.05. The clusters were based on pathway connectivity assessed by the predefined kappa score threshold of 0.4. And the cluster name was represented by the most significant pathway in a cluster.

## Results

### Participant characteristics

In the discovery cohort, the mean age was 58.4 (standard deviation [SD]: 12.2) years and 27.8% were female. The baseline characteristics between individuals with and without CAD endpoints were similar in the discovery set (P ≥ 0.078), except for eGFR (mean: 75.1 ± 17.0 vs. 83.1 ± 14.1, P = 0.038, Table 1). Of 893 participants in the validation cohort, the mean age was 52.7(16.3) years, and 446 (49.9%) were women. Individuals who progressed to CAD endpoints tended to be male, older, having a history of hypertension, diabetes, and lower eGFR, compared to those without CAD endpoints (P ≤ 0.014, Table 2).

**Table 1.**
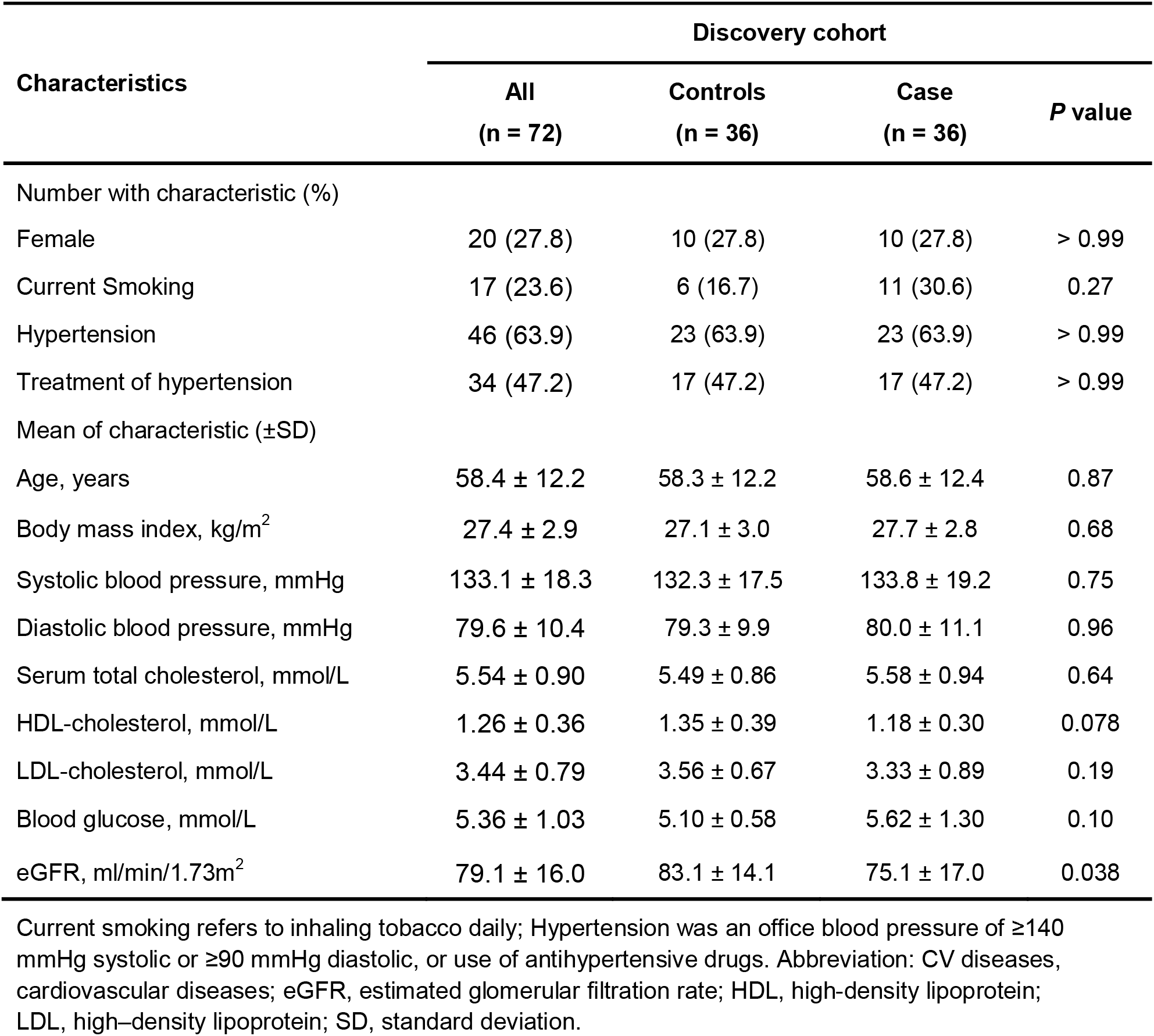
Participant characteristics in the discovery cohort.

| Characteristics | Discovery cohort |  |  | P value |
| --- | --- | --- | --- | --- |
|  | All<br>(n = 72) | Controls<br>(n = 36) | Case<br>(n = 36) |  |
| Number with characteristic (%) |  |  |  |  |
| Female | 20 (27.8) | 10 (27.8) | 10 (27.8) | > 0.99 |
| Current Smoking | 17 (23.6) | 6 (16.7) | 11 (30.6) | 0.27 |
| Hypertension | 46 (63.9) | 23 (63.9) | 23 (63.9) | > 0.99 |
| Treatment of hypertension | 34 (47.2) | 17 (47.2) | 17 (47.2) | > 0.99 |
| Mean of characteristic (±SD) | | | | |
| Age, years | 58.4 ± 12.2 | 58.3 ± 12.2 | 58.6 ± 12.4 | 0.87 |
| Body mass index, kg/m<sup>2</sup> | 27.4 ± 2.9 | 27.1 ± 3.0 | 27.7 ± 2.8 | 0.68 |
| Systolic blood pressure, mmHg | 133.1 ± 18.3 | 132.3 ± 17.5 | 133.8 ± 19.2 | 0.75 |
| Diastolic blood pressure, mmHg | 79.6 ± 10.4 | 79.3 ± 9.9 | 80.0 ± 11.1 | 0.96 |
| Serum total cholesterol, mmol/L | 5.54 ± 0.90 | 5.49 ± 0.86 | 5.58 ± 0.94 | 0.64 |
| HDL-cholesterol, mmol/L | 1.26 ± 0.36 | 1.35 ± 0.39 | 1.18 ± 0.30 | 0.078 |
| LDL-cholesterol, mmol/L | 3.44 ± 0.79 | 3.56 ± 0.67 | 3.33 ± 0.89 | 0.19 |
| Blood glucose, mmol/L | 5.36 ± 1.03 | 5.10 ± 0.58 | 5.62 ± 1.30 | 0.10 |
| eGFR, ml/min/1.73m<sup>2</sup> | 79.1 ± 16.0 | 83.1 ± 14.1 | 75.1 ± 17.0 | 0.038 |

Current smoking refers to inhaling tobacco daily; Hypertension was an office blood pressure of $\geq 140$ mmHg systolic or $\geq 90$ mmHg diastolic, or use of antihypertensive drugs. Abbreviation: CV diseases, cardiovascular diseases; eGFR, estimated glomerular filtration rate; HDL, high-density lipoprotein; LDL, high-density lipoprotein; SD, standard deviation.

**Table 2.** Participant characteristics in the validation cohort.

| Characteristics | Validation cohort |  |  | *P* value |
| --- | --- | --- | --- | --- |
|  | All<br>(n = 893) | Controls<br>(n = 778) | Case<br>(n = 115) |  |
| Female, % | 446 (49.9) | 412 (53.0) | 34 (29.6) | <0.0001 |
| Hypertension, % | 362 (40.5) | 303 (39.0) | 59 (51.3) | 0.014 |
| History of diabetes, % | 117 (13.1) | 65 (8.4) | 52 (45.2) | <0.0001 |
| Age, years | 52.7 ± 16.3 | 50.8 ± 15.9 | 65.6 ± 12.9 | <0.0001 |
| Systolic blood pressure, mmHg | 130.4 ± 18.1 | 129.2 ± 17.5 | 138.4 ± 20.0 | <0.0001 |
| Diastolic blood pressure, mmHg | 79.2 ± 9.7 | 79.4 ± 9.6 | 77.9 ± 10.7 | 0.31 |
| Mean arterial pressure, mmHg | 109.8 ± 14.5 | 110.6 ± 14.0 | 104.2 ± 16.9 | <0.0001 |
| eGFR, ml/min/1.73m<sup>2</sup> | 81.1 ± 22.6 | 83.9 ± 20.8 | 62.1 ± 25.3 | <0.0001 |

Hypertension was an office blood pressure of ≥140 mmHg systolic or ≥90 mmHg diastolic, or use of antihypertensive drugs. Abbreviation: eGFR, estimated glomerular filtration rate.

### Urinary proteomic classifiers and the prediction of CAD

In the discovery cohort, a total of 518 sequenced peptides significantly differed between individuals with and without CAD after multiple testing corrections and permutation tests.

Subsequently, a urinary proteomic classifier integrating 160 significant peptides was developed in the discovery cohort using the SVM (regularization parameter: 64, kernel coefficient: 0.000256, epsilon: 0.001). When generalizing the new classifier to the independent validation cohort, this new classifier provided an AUC of 0.77 (95%CI: 0.70-0.84), 0.83 (95%CI: 0.76-0.86), and 0.82 (95%CI: 0.78-0.87) for the prediction of CAD endpoints at 3, 5, and 8-year, consistently outperforming to prior classifiers, CAD238 and ACSP75 (Table 3). Moreover, the addition of the new classifier significantly improved AUC on top of CAD238 (from 0.65-0.71 to 0.79-0.85) and ACSP75 (from 0.53-0.61 to 0.77-0.84). The combination of three classifiers yielded an increased AUC of 0.79 (0.73-0.85), 0.85 (0.78-0.87), and 0.84 (0.80-0.89) for the prediction of 3, 5, and 8-year CAD. Furthermore, the risk reclassification was improved by the addition of the new classifier on top of the Framingham risk score in 737 individuals from the FLEMENGHO study, as suggested by an NRI of 0.61 (95% CI: 0.25-0.95, P = 0.001) and IDI of 0.02 (95% CI: 0.02-0.05, P = 0.39).

**Table 3.** Predictive performance of urinary proteomic markers for CAD.

|  | Time dependent AUC (95% CI) |  |  |
| --- | --- | --- | --- |
|  | 3-year | 5-year | 8-year |
| **No. events/ at risk** | **69/784** | **102/716** | **111/478** |
| 160-marker | 0.77 (0.70-0.84) | 0.83 (0.76-0.86) | 0.82 (0.78-0.87) |
| CAD238 | 0.65 (0.58-0.72) | 0.66 (0.61-0.73) | 0.71 (0.66-0.77) |
| ACSP75 | 0.53 (0.45-0.61) | 0.61 (0.48-0.64) | 0.53 (0.47-0.60) |
| 160-marker+CAD238 | 0.79 (0.72-0.86) * | 0.84 (0.77-0.88) * | 0.85 (0.80-0.89) * |
| 160-marker+ACSP75 | 0.77 (0.71-0.84) † | 0.84 (0.77-0.86) † | 0.82 (0.78-0.86) † |
| 160-marker+CAD238+ACSP75 | 0.79 (0.73-0.85) | 0.85 (0.78-0.87) | 0.84 (0.80-0.89) ‡ |

\* AUC was significantly improved (P <0.05) compared with the model with CAD238.
† AUC was significantly improved (P <0.05) compared with the model with ACSP75.
‡ AUC was significantly improved (P <0.05) compared with the model with 160-marker.

### Association between urinary proteomic classifier with CAD

The association between the new classifier and the risk of CAD is presented in Table 4. In the multivariable-adjusted model, a 1-SD increment in the new classifier was associated with a 1.62-fold (95% CI: 1.34-1.96) higher risk of incident CAD at 8-year follow-up. Compared with individuals in the bottom quartile of the new classifier, those in the highest quartile had a significantly higher risk of CAD (HR: 3.55, 95% CI: 1.84-6.85) after adjustment. When additionally adjusting for eGFR, slightly weakened associations that remained significant (HR: 1.54, 95% CI: 1.26-1.89 for 1-SD increment) were observed for the new classifier. Neither CAD238 nor ACSP75 was significantly associated with CAD after adjustment. Moreover, for 737 individuals from the FLEMENGHO study, the new classifier was positively associated with CAD (adjusted HR: 1.44, 95% CI: 1.01-2.95 for 1-SD increment), independent of the Framingham risk score.

**Table 4.** Risk of CAD by baseline urinary proteomic classifier.

|  | Events/at risk | Multivariable model 1 |  | Multivariable model 2 |  |
| --- | --- | --- | --- | --- | --- |
|  |  | HR (95% CI) | P | HR (95% CI) | P |
| 160-marker |  |  |  |  |  |
| Per 1-SD increment | 115/893 | 1.62 (1.34-1.96) | <0.0001 | 1.54 (1.26-1.89) | <0.0001 |
| Quartile 1 | 11/223 | Reference |  | Reference |  |
| Quartile 2 | 9/223 | 0.89 (0.37-2.17) | 0.80 | 0.88 (0.36-2.14) | 0.78 |
| Quartile 3 | 26/225 | 2.27 (1.11-4.62) | 0.024 | 2.14 (1.05-4.38) | 0.037 |
| Quartile 4 | 69/222 | 3.55 (1.84-6.85) | 0.0002 | 3.19 (1.63-6.27) | 0.0007 |
| CAD238 |  |  |  |  |  |
| Per 1-SD increment | 115/893 | 1.03 (0.86-1.22) | 0.78 | 1.01 (0.85-1.21) | 0.90 |
| Quartile 1 | 13/223 | Reference |  | Reference |  |
| Quartile 2 | 27/224 | 2.35 (1.21-4.57) | 0.012 | 2.25 (1.16-4.39) | 0.017 |
| Quartile 3 | 25/222 | 1.35 (0.68-2.66) | 0.39 | 1.19 (0.60-2.38) | 0.61 |
| Quartile 4 | 50/224 | 1.64 (0.85-3.15) | 0.14 | 1.47 (0.76-2.84) | 0.25 |
| ACSP75 |  |  |  |  |  |
| Per 1-SD increment | 115/893 | 1.05 (0.87-1.28) | 0.61 | 1.09 (0.90-1.32) | 0.38 |
| Quartile 1 | 29/223 | Reference |  | Reference |  |
| Quartile 2 | 20/223 | 0.51 (0.29-0.90) | 0.021 | 0.55 (0.31-0.97) | 0.040 |
| Quartile 3 | 26/224 | 0.76 (0.44-1.29) | 0.30 | 0.79 (0.46-1.35) | 0.39 |
| Quartile 4 | 40/223 | 0.98 (0.61-1.59) | 0.94 | 1.09 (0.67-1.77) | 0.74 |

Model 1 was adjusted for sex, age, mean arterial blood pressure, and history of diabetes. Model 2 was adjusted as for model 1 and for eGFR.

### Urinary peptides associated with CAD

These 160 urinary peptides were fragments of 58 parental proteins, mainly derived from collagens, such as collagen type I alpha I chain (36 peptide, 22.5%), collagen type III alpha chain (22 peptide, 13.8%), and collagen type I alpha II chain (9 peptides, 5.6%). Of 160 urinary peptides, most peptides were higher in individuals with CAD, except for 27 peptides from collagen type I alpha I chain (7 peptides), collagen type III alpha chain (5 peptides), uromodulin (6 peptides), apolipoprotein C-II (1 peptide), sarcalumenin (1 peptide), collagen type 1 alpha I (1 peptide), collagen type III alpha 1 (2 peptides), and collagen V alpha 2 (1 peptide). Table S1 in the supplementary materials displays the list of 160 peptides, fold-change in individuals with CAD, and their parental proteins. The elevated excretion of most collagen I and III fragments in individuals with CAD might suggest an upregulated collagen degradation. In addition to collagens, other prominent proteins, including fibrinogen (10 peptides), uromodulin (7 peptides), polymeric immunoglobulin receptor (3 peptides), CD99 antigen (2 peptides), and insulin (2 peptides), and insulin-like growth factors II were upregulated in participants with CAD. The remaining parental proteins functioned diversely, including lipid metabolism (apolipoprotein C-II, apolipoprotein C-III, apolipoprotein L1, clusterin), proliferation (thymosin beta-4), tissue remodeling (clusterin), immune response (complement C4-A, complement factor B), inflammation (interleukin-1 receptor antagonist protein).

Bioinformatics was performed to obtain a comprehensive overview of the proteases that could lead to the generation of peptides associated with CAD. According to the cleavage sites, 11 proteases were predicted to participate in the fragmentation of proteins associated with CAD. Peptides that were higher in individuals with CAD predicted 11 up-regulated matrix metalloproteinases (MMPs) that mainly involve in collagen degradation, including MMP 1, 2, 8, 9, 13, 14, cathepsin K, and neuroendocrine convertase 1. Down-regulated peptides predicted lower activity of MMP 2 and 9 as well. (Table S2 in the supplementary materials).

The parental proteins of urinary peptides and their upstream proteases might be involved in the pathogenesis of CAD. Pathway enrichment analysis mapped 8 clusters consisting of 41 pathways (Figure 2 and Table S3 in the supplementary materials). The major pathways enriched were extracellular matrix turnover and signaling, cell surface interactions, plasma lipoprotein metabolism, activation of MMP, complement cascade, proliferation, and insulin processing.

**Figure 2.**
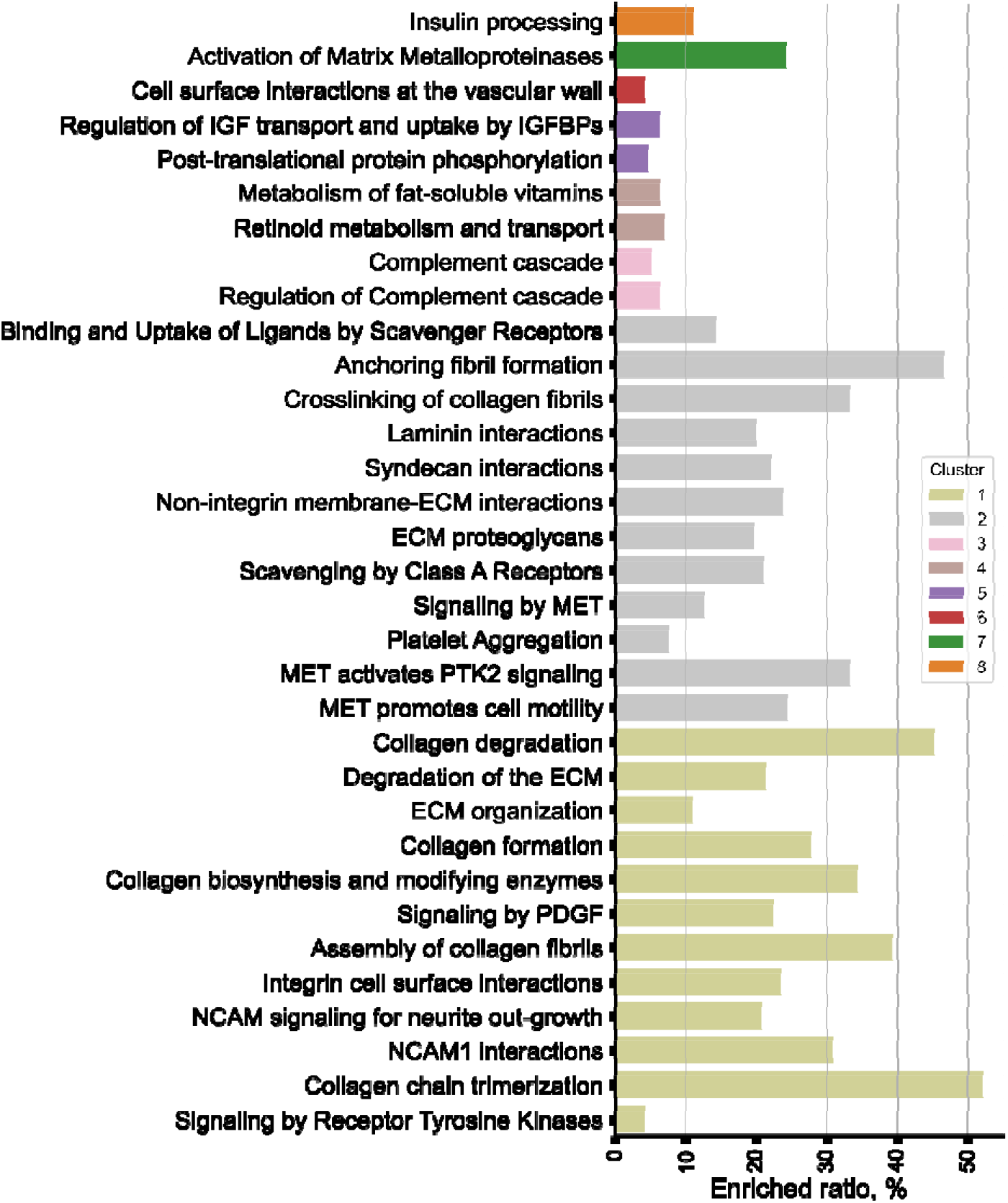
Pathway enrichment analysis based on the parental proteins and predicted proteases of 160 urinary peptides. Enriched pathways involved in seven clusters: extracellular matrix turnover and signaling (cluster 1), cell surface interactions at the vascular wall (cluster 2), plasma lipoprotein assembly, remodeling, and clearance (cluster 3), activation of matrix metalloproteinases (cluster 4), complement cascade (cluster 5), regulation of IGF transport and uptake by IGFBPs (cluster 6), insulin processing (cluster 7). All displayed pathways had P-value < 0.05. Enrichment ratio represents the number of submitted proteins in a particular pathway to the total number of proteins in the pathway. ECM, extracellular matrix; LPL, lipoprotein lipase; LIPC, hepatic lipase; MET, receptor tyrosine kinase; NCAM1, neural cell adhesion molecule. NGF, nerve growth factor; PDGF, platelet-derived growth factor; PTK, protein tyrosine kinase; IGF, insulin-like growth factor; IGFBP, Insulin-like growth factor binding protein.

## Discussion

In the present study, we developed a novel urinary proteomic classifier associated with CAD using a machine learning approach and validated its predictive performance in an independent prospective cohort of 893 individuals. Specifically, the novel proteomic classifier outperformed previously developed CAD-related classifiers in terms of the prediction of CAD endpoints, and the addition of the novel proteomic classifier to previous classifiers significantly improved its predictive performance, with the highest AUC of 0.87. Moreover, using multivariable Cox regression models, the novel proteomic classifier was significantly associated with the risk of reaching an ischemic endpoint at 8-year follow up. The novel proteomic classifier was comprised of 160 urinary polypeptides that may provide mechanistic insights into CAD and suggest targeted interventions to halt or reverse the development and progression of ischemic heart disease.

Accumulating evidence indicated that urinary proteome analysis identifies biomarkers for diverse disease conditions, including CAD. Zimmerli *et al*. previously analyzed urinary proteomics in 88 CAD patients and 282 controls.^26^ A panel consisting of 15 urinary polypeptides was constructed and provided a sensitivity of 98% and specificity of 83% for the detection of CAD.^26^ Subsequently, another urinary proteomic signature was developed with 17 peptides in 15 CAD patients and 14 controls and yielded a sensitivity of 81% and specificity of 92% in an independent case-control set (22 CAD cases and 16 controls).^27^ However, the diagnostic performance of these two urinary panels declined, as reflected by an AUC of 0.68 and 0.77, respectively, when extending to an independent set with 71 CAD patients and 67 controls.^10^ Given the declined performance, Delles *et al* further developed and validated a urinary proteomic pattern with 238 polypeptides, CAD238, for the diagnosis of CAD in 408 participants using the support vector machine.^10^ CAD238 showed an AUC of 0.87 with a sensitivity of 79% and specificity of 88%.^10^ CAD238 was suggested to be associated with CAD.^11^ Likewise, a urinary proteomic pattern based on 75 polypeptides (ACSP75) was proposed for the prediction of acute coronary syndromes.^13^ There is room for improvement as suggested by the AUC (0.64) of ACSP75 in a validation set of 42 cases and 42 controls. While these previous findings clearly support the concept and utilization of urinary proteome analysis for CAD-specific biomarker discovery, we set out to further improve its predictive performance in a relatively large and well characterized cohort.

It is challenging to diagnose and intervene in CAD in the early stage, as CAD is characterized by coronary artery atherosclerosis that can progress asymptomatically for extended periods of time. Therefore, prognostic biomarkers are important to stratify CAD risk and identify individuals with high risk who might benefit from early interventions. Our findings show that the novel proteomic classifier was associated with CAD risk, superior to CAD238 and ASCP75, in a validation cohort that included participants with various clinical contexts and hence risk profiles from the general population to patients with diabetes. This attempt enables a robust evaluation of the prognostic value of the new proteomic biomarker. It would allow the proteomic classifier to retain its prognostic discrimination when generalizing it to other cohorts.

Despite the advances in urinary proteomic biomarker research in the context of CAD, several hurdles remain. First, it is unclear whether the novel urinary proteomic classifier could distinguish the subtypes of CAD, such as acute myocardial infarction, angina, and cardiac arrest. The risk assessment of CAD subtypes would stimulate individualized prevention and treatment strategies that would be more effective than a one-size-fits-all strategy. The existing biomarkers were specifically developed to detect composite CAD endpoints or a single subtype, such as ACSP75. Second, the developed CAD-specific biomarkers require external validation, especially in cohorts with distinct comorbidities. Oellgaard et al. reported that ACSP75 was associated with cardiovascular events, but CAD238 did not present a significant prognostic association in patients with diabetes.^28^ Although this novel proteomic classifier showed prognostic potential for individuals, including diabetes patients, large, multicentre prospective studies may help to resolve these challenges.

Endogenous peptides and small proteins in urine provide clues to the status of their larger precursor proteins, including the degradation process and post-translational modifications.^6^ These subtle clues can help to puzzle out complex pathological mechanisms. The pathogenesis of CAD is complex and multifactorial, caused by diverse mechanisms. The complex nature of CAD that is hardly reflected by a single molecule requires a comprehensive search and integration of biomarkers. Previous urinary proteomic panels proposed by Zimmerli et al and Von Zur Muhlen et al are comprised mainly of collagen I and collagen type III fragments.^26, 27^ The constitution of CAD238 and ACSP75 is more diverse, involving collagen, fibrinogen, and mucin.^10, 13^ It appears that collagen turnover in the extracellular matrix is disproportionally regulated in individuals prior to their CAD events. Collagen type I contributes to 60% of protein contents in the atherosclerotic plaque, and is an essential component of the fibrous cap.^29^ Upregulated degradation of collagen type I can lead to a thinner fibrous cap, a trigger for atherosclerotic plaque rupture.^30^ Circulating MMP-mediated collagen type I biomarker has been suggested to be positively associated with a higher risk of cardiovascular events.^29^ Our study suggest that the majority of collagen type I and III fragments were more abundantly excreted in urine, prior to future CAD events, indicating an upregulated collagen degradation in extracellular matrix. This was consistent with the predicted upregulated MMPs, such as MMP 1, 2, 8, 9, 13, 14, and cathepsin K.

The overlap between our proteomic characterization, CAD238, and ACSP75 also included uromodulin. Uromodulin, produced by renal epithelial cells, is considered the most abundant glycoprotein in urine and can be found in the blood as well.^31^ Uromodulin is frequently associated with the risk of chronic kidney disease and hypertension.^31, 32^ Recent evidence demonstrated a reverse association of serum uromodulin with coronary artery events and coronary artery calcification, even though the mechanism is unclear.^33, 34^ Similarly, we also observed the level of urinary uromodulin peptides was lower in individuals who progressed to CAD.

There were several unique proteins included in the novel proteomic classifier. For instance, clusterin is a glycoprotein, also known as apolipoprotein J that plays an essential role in inflammation, lipids metabolism, and atherosclerosis.^35^ High serum clusterin is suggested to be associated with premature coronary artery disease.^36^ Consistent with this, we also observed a higher urinary clusterin peptide for individuals with CAD endpoints. Carbonic anhydrase I is an enzyme that catalyzes the reversible hydration of carbon dioxide, and it links to atherosclerotic calcification and the progression of atherosclerosis.^37^ Folate receptor alpha CD99 is another significantly down-regulated protein in our urinary proteomic characterization. CD99, a cell adhesion molecule, is expressed by endothelial cells and engages in the recruitment of monocytes and lymphocytes. The recruited inflammatory cells can migrate to atherosclerotic regions and initial inflammation.^38^ Vaccination against CD99 can effectively attenuate the progression of atherosclerotic plaques by lowering leukocytes in atherosclerotic lesions.^39^

### Strengths and limitations

Our study has several strengths, including systematic urinary proteome analyses with good reproducibility, validation of the urinary proteomic in an independent, relatively large cohort, and assessment of the prognostic value. There are several limitations of our study. First, the association between the urinary proteomic classifier and the severity or subtypes of coronary events was not investigated, which requires a large, prospective study. Second, future studies are warranted to further evaluate whether the urinary proteomic classifier can identify plaque volume and composition measured by ultrasound or computerized tomography. Correlating urinary biomarkers with subclinical lesions could help to determine when interventions, such as statin therapy, should be initiated. Last, pathway analysis of these peptides relied on existing studies, thus their roles in the development of CAD might be different, which needs to be determined in preclinical experimental and epidemiological studies.

## Conclusions

This study developed and validated a urinary proteomic classifier for the prediction of ischemic CAD endpoint with good performance. The peptides constituting the proteomic classifier were involved in diverse pathways associated with atherosclerosis, including collagen turnovers, lipid metabolism, and inflammation.

## Data Availability

The data underlying this article cannot be shared publicly due to the privacy of the participants. The data can only be reasonably requested to the corresponding author

## Acknowledgements

The authors gratefully acknowledge the contribution of Prof. Harald Mischak and Agnieszka Latosinska from Mosaiques Diagnostics (Hannover, Germany) for urinary proteomic profiling, data analyses, and interpretation and the clerical contribution of Renilde Wolfs.

## Authors’ Contributions

D.M. W and Z.-Y. Z. conceptualized and designed the study. Z.-Y.Z. contributed to data acquisition. D.M. W, and Z.-Y. Z. performed analysis. D.M. W initially drafted the manuscript. All authors interpreted the data and critically revised the manuscript. All authors reviewed and approved the final manuscript.

## Supplementary materials

Tables S1–3

